# How ready is the health care system in North East India for surgical delivery? A mixed methods study on surgical capacity and need

**DOI:** 10.1101/2023.06.19.23291617

**Authors:** Amrit Virk, Rebecca King, Michael Heneise, Julia Brown, David Jayne, Tim Ensor

## Abstract

**Background:** Surgical services are scarce with persisting inequalities in access across populations and regions globally. As the world’s most populous county, India’s surgical need is high and delivery rates estimated to be sub-par to meet need. There is a dearth of evidence particularly sub-regional data on surgical provisioning and need to aid planning.

**Aim and method:** This mixed methods study examines the state of surgical care in Northeast India, specifically health care system capacity and barriers to surgical delivery. It involved a facility based census and semi-structured interviews with surgeons and patients across four states in the region.

**Results:** Abdominal conditions constituted a large portion of the overall surgeries across public and private facilities in the region. Workloads varied among surgical providers across facilities. Task-shifting occurred, involving non-specialist nursing staff assisting doctors on surgical procedures or surgeons’ taking on anaesthetic tasks. Structural factors dis-incentivised facility level investment in suitable infrastructure. Patients’ care pathways were shaped by facility level shortages as well as personal preferences influenced by cost and distance to facilities.

**Discussion and conclusion:** skewed workloads across facilities and regions indicate uneven surgical delivery, with potentially variable care quality and provider efficiency. A systemic approach to referral coordination and human resource management are evident. Existing task shifting practices, along with incapacities induced by structural factors signal the direction of possible policy action.

## Introduction

Conditions treatable by surgery are nearly a third of the global disease burden making it a critical area of public health and policy attention (Shrime et al 2015). The investment case for surgery is evident— essential surgical procedures and anaesthesia, most of which could be provided at first-level hospitals, are a cost-effective undertaking with substantial returns on investment, preventing as many as 6-7% of all avoidable deaths in low and middle income countries(LMICs) (Mock et al 2015). Yet, a staggering 5 billion people – approximately 5 of every 7 people globally – lack access to safe, timely, and affordable surgical care (Jamison et al 2013; Debas et al 2015). Moreover, fewer than 6% of all surgical operations occur in LMICs where over a third of the world’s population lives (Meara et al 2105). The figures reveal vast geographical disparities, 295 in LMICs against 23,000 surgeries per 100,000 population in high-income countries, driven by shortages of medical staff, poor access to healthcare services, and weak record keeping (Bhandarkar et al 2021). Hence, surgical resources need to be urgently scaled-up in pursuit of achieving universal health coverage (UHC), to ensure everyone receives good quality and affordable health care services that they need (WHO 2005). Three procedures, caesarean section, fracture repair, and laparotomy together known as the Bellwether Procedures can treat a vast majority of surgical problems; ensuring they are available 24/7 at all first level hospitals a key strategy to expand surgical access (O’Neill 2016).

Approximations of surgical care in India indicate substantial gaps in meeting required needs; a recent study estimating a requirement for 3646 surgeries annually per 100,000 population (Bhandarkar et al 2021). Astonishingly, as many as 90% of people in rural India lack access to safe surgical care (LCoGSC 2019), and current rates of surgical procedures being far below global averages (50–499 surgeries per 100,000) (Singh et al 2022). Within India, the health system in the North-Eastern states is among the most underdeveloped with poor access and health indicators (Ensor et al 2020). This region is also distinct in Eurasia, characterised by ethnolinguistic diversity, indicating that many vernacular health systems interact (Post et al 2022). Moreover, at both health system and community levels, a lot remains unknown about the structures, processes, and practices for surgical delivery and care seeking in rural and NE India. System readiness and potential to provide basic surgical care is also vital to inform technological and policy interventions to improve surgical care for rural areas of India.

### Aim

This mixed methods study aimed to examine the state of surgical care in North Eastern India, namely health care system capacity and barriers to surgical care. It thereby develops a baseline for understanding the readiness and potential of facilities to deliver surgical care. This study addresses a vital gap in current literature (Zadey et al 2023), by providing sub-national evidence on the current state of surgical care in India.

## Materials and Methods

### Study design

This mixed methods study involved qualitative interviews and health facility surveys conducted across four (of eight) states in NE India: Arunachal Pradesh, Assam, Nagaland, and Manipur between April 2019 and May 2020, together representative of a substantive geographic area. The facility survey was designed to assess healthcare system readiness to provide basic surgical care and abdominal surgery. Semi-structured interviews focused further on surgical provisioning and patient care seeking behaviours to uncover barriers to surgical delivery and patterns of care seeking. A preliminary review of relevant secondary literature, national plans, and country reports accompanied this assessment of the existing state of surgical care in NE India.

The survey tool (box 1) and interview guides were drafted following a series of meetings among a multi-disciplinary team based in the UK and India (surgeons, economists, anthropologists, statisticians, and social scientists). Interview tools, including topic guides were prepared in both English and Hindi, and back translated to check for accuracy. The local team conducted a few interviews in Nagamese (Nagaland), Assamese (Assam), and Meitei (Manipur), consulting a translator for Meitei. The survey tool and topic guides for each respondent category were further refined after initial pilot-testing in the field.

### Site selection

Convenience and purposive site selection led to the focus on four study districts (Table 1) where local doctors trained in a novel surgical technique by members of the research team came from the NIHR Global Health Research Group (Panel 1). Local informants with long-standing clinical and research knowledge of the region input into site selection to also ensure a comparison between larger, urban districts (Dimapur) and rural locales. The facility survey was a census of all surgical facilities in these four districts and interviewees were also located here. Supported by UK and India based senior researchers, a 7-member team of local research assistants carried out the facility surveys and interviews between May 2019-January 2020. For the latter, study methods are reported based on guidance adapted from Standards for reporting qualitative research (SRQR) guidelines.^13^

**Table 1:** Sample of respondents

| State | District | No. of doctors interviewed | No. of patients interviewed |
| --- | --- | --- | --- |
| Arunachal Pradesh | Bomdila/Bhaloukpong | 1 | 0 |
| Assam | Chirang/Bongaigaon | 2 | 3 |
| Manipur | Churachandpur | 3 | 4 |
| Nagaland | Dimapur | 2 | 4 |
| Total |  | 8 | 11 |

#### Panel 1: NIHR Global Health Research Group-Surgical Technologies

This study is embedded within a large, multi-disciplinary project involving social science, engineering, statistical, and clinical expertise to develop low-cost surgical technology to address unmet surgical need in under-resourced settings in North East India. The project focused on abdominal surgery as a significant component of overall surgical demand in the country. A mixed­ methods study was designed to generate contextual information on health system capacity and care seeking behaviours to inform the work of the other components to develop relevant clinical and engineering interventions.The UK based team worked in close collaboration with regional institutions, including a clinician-led, humanitarian organization providing medical care and surgical training in the region and a team of social scientists from the region.

### Semi-structured interviews

This paper reports on interviews with 8 clinicians and 11 patients across 4 states (Table 1). Participants at surveyed facilities were recruited between May-November 2019 through purposive and snowballing methods to include those with first-hand knowledge and experience of surgical delivery and use. (Table 1). Most doctors (6) worked in private facilities including one at a charitable/mission hospital. Two government doctors in the sample, both from Manipur, also worked at other facilities. All patients interviewed were admitted in private facilities and most (9) were women. Participants were mostly between 30 and 45, with one male in his twenties and an older female patient who was 52 (Table 2).

**Table 2:** profile of patients interviewed

|  |  |  |
| --- | --- | --- |
| Female, 28-35 | Bongaigaon | Assam |
| Female, 30-35 | Bongaigaon | Assam |
| Male, 40-45 | Chirang | Assam |
| Female, 40-45 | Churachandpur | Manipur |
| Female, 52 | Churachandpur | Manipur |
| Female, 35-40 | Churachandpur | Manipur |
| Female, 30-35 | Churachandpur | Manipur |
| Female, 34-40 | Dimapur | Nagaland |
| Female, 44 | Dimapur | Nagaland |
| Male, 22-25 | Dimapur | Nagaland |
| Male, 30-35 | Dimapur | Nagaland |

Local researchers translated any oral or written communication from interviewees that was not in English. An information sheet detailing the purpose of the study, confidentiality and anonymity clauses, and the implications of their involvement was shared in advance with all participants. Informed written / recorded consent from all participants preceded data collection activities. All interviews were audio recorded, transcribed into English, and analysed using NVivo V.12.

All data was anonymised by the local team in India and authors did not have access to information that could identify individual participants during or after data collection. Data analysis involved a deductive aspect given the broader focus on health system capacity and user need as well as a critical inductive dimension wherein themes were extracted from reading gathered data sources. Qualitative data was analysed iteratively, with data gathering and analysis occurring in tandem, and broadly guided by the framework analysis approach^13^.In practice, this involved regular de-briefs within the local team which kept a field diary recording personal reflections. Additionally, the UK and India researchers teleconferenced at regular intervals to discuss emerging themes and connections within the data. The local team transcribed interview data, and a single UK based researcher (AV) then input and coded data using NVivo v12 (QSR 2018). The UK and India lead research leads (AV, MH) also met to discuss and clarify initial codes and evolving themes.

### Facility survey

To conduct the facility census, the team contacted the administrative departments of all listed hospitals in the districts requesting access to logbooks and surgical records on condition of anonymity. Data were collected on a range of variables including monthly patient numbers, surgeries, access (Ref. Panel 2; for e.g. how far patients had to travel, means of transport), size of population served by facility, rural-urban loads, and number of abdominal surgeries annually. The data was digitized on the spot as the log book/ surgical record books could not be removed from the hospital.

Survey data was entered into an appropriate database and then converted to Stata for analysis. Data entry was double-entry and undertaken by the local research team. Data had no personal identifier so was anonymous at the individual level. Facility level identifiers were included to facilitate later linkage to other data sets using geographic coding.

The results below summarise the information from the quantitative and qualitative data to present a composite analysis of surgical capacity and challenges to effective provisioning of surgical care.

#### Panel 2: Facility functionality

The functionality (F) of facilities was captured across six domains:

i. availability of a surgeon 24/7 in the facilities D_1_[0,1], taking a value of 1 if a surgeon is available 24 hours a day and reduced proportionatelyfor less availability;
ii. availability of basic infrastructure D_2_[0,1], taking a value of 1 where electricity, water and oxygen are available 24 hours a day and reduced by a 1/3 for each utility not available;
iii. ability to provide blood transfusions D_3_[0,1], taking a value of 1 if blood­banking and transfusion equipment are available plus ability to cross-match, reduced by 1/3 for each component lacking;
iv. availability of an anaesthetics D_4_[0,1], taking a value of 1 if an anaesthetist (specialist, doctor or trained non-doctor) and anaesthetic equipment are available, reduced by½ for each component lacking;
v. ability to undertake basic lab tests D_5_[0,1], taking a value of 1 where all essential tests are available (complete blood count, pregnancy, coagulation, urine analysis and infectious panel of tests) and reduced by 1/5 for each component lacking; and
vi. availability of basic surgical equipment D_6_[0,1], taking a value of 1 where all 44 items are available, reduced by1/44for each item lacking.

Each of these domains were scored and given a weighting of 1/6th in a total index of functionality to deliver surgical services, i.e:

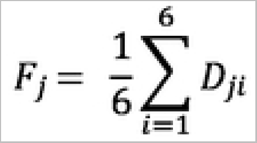

## Results

### Overview: Range of surgical procedures and use

Both, three-month surgical logbooks and aggregate annual reports provide a similar picture of the main types of surgery provided across the region. Caesarean section (offered by 18/19 facilities providing surgical data), cholecystectomy (12/19), and appendectomy (14/19) constitute 63% of surgery in all facilities and 76% in government facilities (Table 3). A small number of other services, largely provided in non-government facilities, included hysterectomy, laparotomy, surgical repair of fractures and hernias, making up a further 30% of surgeries. The remaining 6% is made up of more than 20 infrequently provided surgical services. According to the aggregate reports, the bellwether procedures (Caesarean section, laparotomy and fracture fixation) account for 45% of all surgery in all facilities across the region. The survey showed that although open surgery remains the dominant technique, laparoscopic techniques are increasingly common particularly in private facilities with around 28% of laparoscopic-amenable surgery carried out in this way.

**Table 3:**
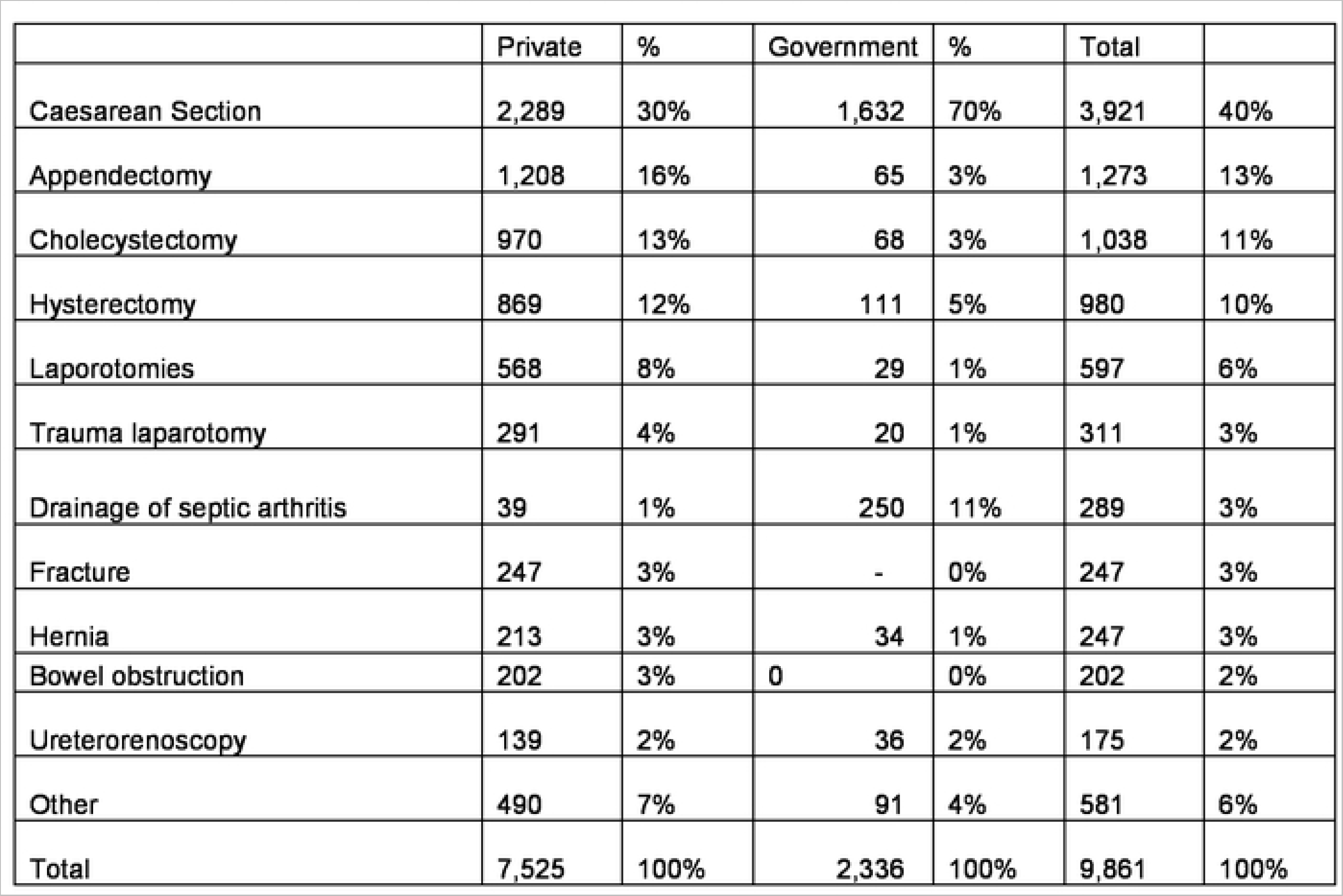
Type of surgery by provider ownership (based on annual reporting)

*Gender and surgery:* The surgical logbooks suggest that women undergoing surgery out-number men by more than 3 to 1. This ratio falls to 1.4 if caesarean sections and hysterectomy are excluded, highlighting the prominence of these two obstetric procedures in the region. In the facilities surveyed, women outnumber men for most procedures even those that are typically emergencies such as appendectomy. The median age for men undergoing surgery is 39 (Interquartile range 30) while for women it is 34, 36 (IQR 19) when caesarean sections are excluded. There is an even split between surgery carried out under general anaesthetic (47%) and under spinal anaesthesia (46%) with the remainder undertaken using local sedation such as ketamine. If caesarean sections are excluded, more than 70% of surgery is undertaken using general anaesthetic.

**Facility functionality for delivery:** Based on the 6-domain index of functionality, overall functionality appeared to be a little higher in non-government (median 0.86IQR 0.19) compared to government (median 0.7, IQR 0.37) facilities. Public facilities were more likely to have facilities for anaesthesia in place while private facilities appear more likely to provide 24-hour surgical cover, blood banking and available surgical equipment (Figure 1).

**Figure 1:**
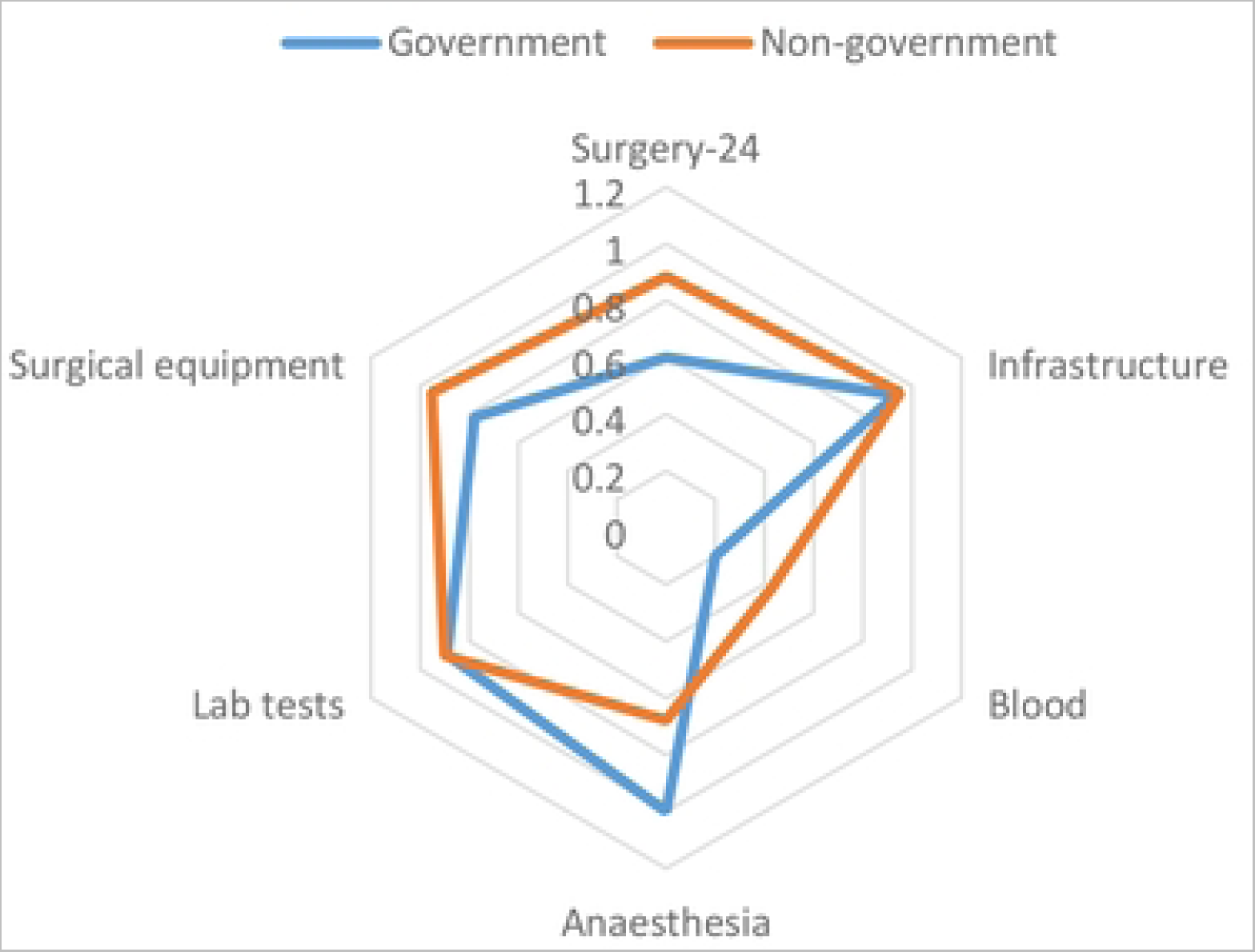
Domains of facility functionality for government and non-government facilities

A positive relationship between facility functionality and surgical volume was evident (Figure 2), reflecting the importance of infrastructure to service provision.

**Figure 2:**
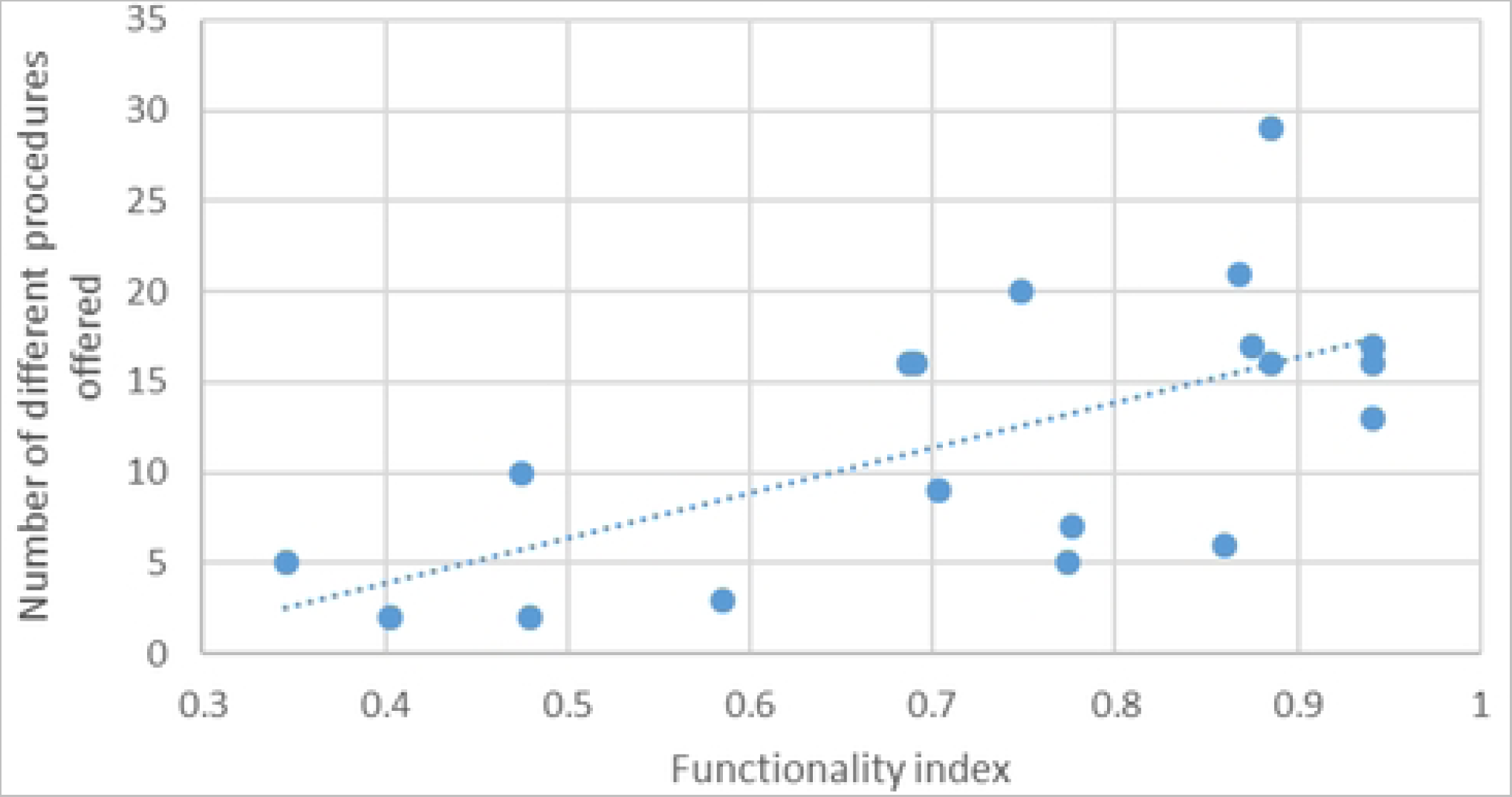
Number different surgical procedures and facility functionality

In general, private facilities appear to offer a wider range of procedures, an average of 14 out of the 34 listed in the survey instrument (Table 3). In contrast, government facilities offer 5-6 procedures on average; most commonly: caesarean section, appendectomy, hysterectomy, tubal ligation, laparotomy, and cholecystectomy. Although these results will need to be read with caution as there were significantly fewer (n=5) public facilities among the universe of surveyed facilities. There appears to be a positive association between the number of different surgical procedures provided by a facility and the functionality of that facility (Figure 2).

The assessment suggested that most facilities have adequate infrastructure, ability to undertake essential laboratory tests, and surgical equipment. Most facilities could provide near 24-hour access to X-ray machines, although access to other scanning technology is much lower particularly in rural areas (see figure 3). Interviews with doctors similarly revealed use of old equipment for surgeries, and deficiencies in availability and quality. In some cases, nursing staff were relied on to carry out regular maintenance such as oiling hand-held equipment but the nursing staff were not in a position to do this for electrical equipment that needed trained technicians. The following quote from a government facility illustrates this,

**Figure 3:**
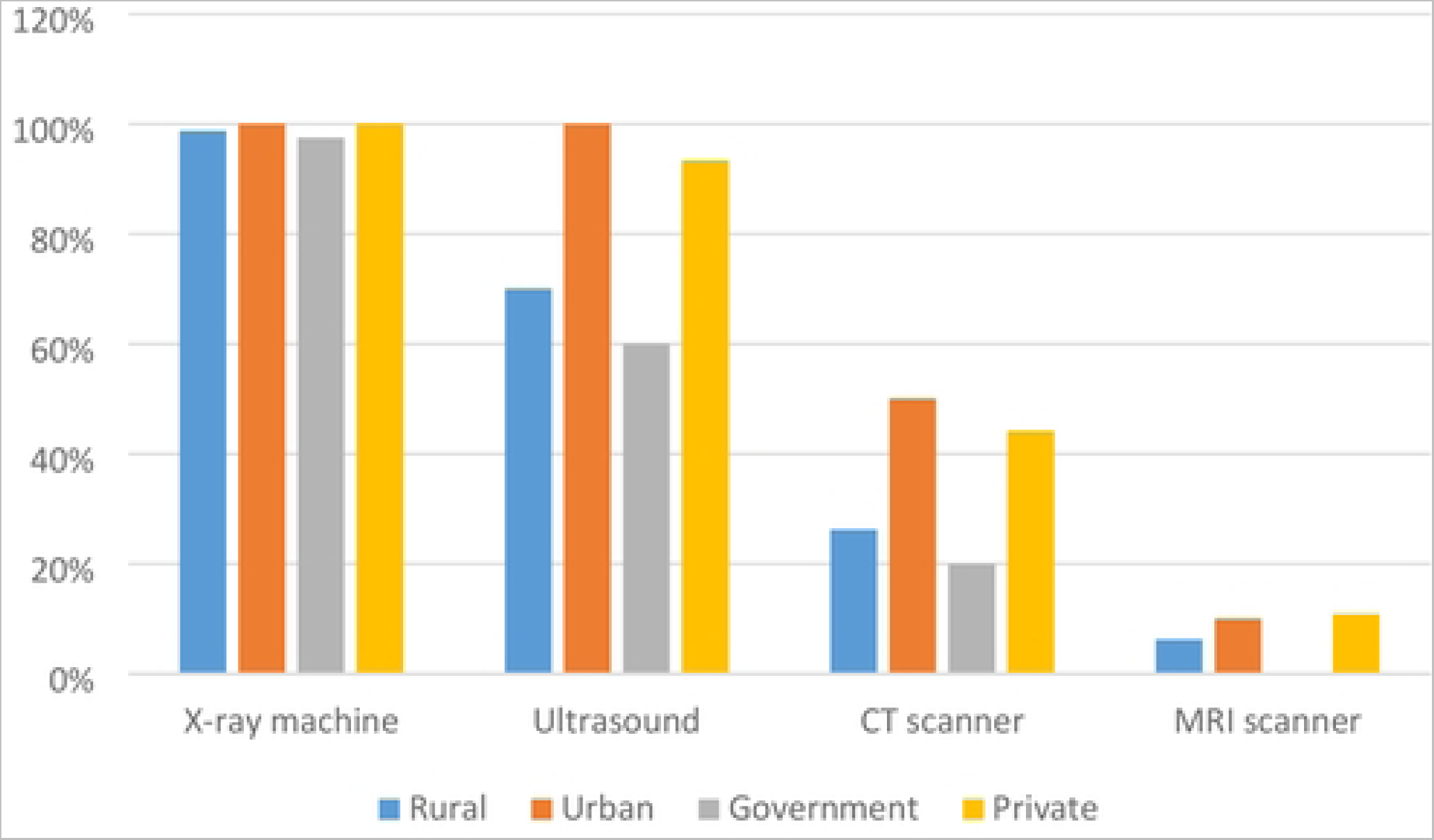
Availability of scanning technology

*CT scan we don’t have… we had CT scan but then once it broke down after that nobody is repairing it. The government is also not repairing it. But other than that, like Ultrasound is free of cost, colonoscopy is also free of cost, x-ray also.*(Govt doctor, Manipur, MCPT002MP)

### Surgical workloads and capacity

All surveyed facilities had surgeons available 24 hours a day, yet most facilities will often have a single surgeon on shift at any one time. Survey results indicated relative paucity of surgeons and anaesthetists in some facilities contributing to huge variation in surgical caseloads. The survey found surgical workloads varying from 25 to almost 3,000 operations per year (Table 4). The narrower range of procedures offered at government facilities mean that while overall surgical loads appear to be higher in non-government facilities, the average number of operations of each procedure are considerably lower in such facilities.

**Table 4:** Surgical workload

|  | Surgeons | Anaesthetists | Average number distinct procedures undertaken | Operations per facility | Average operations per different procedure | Operations per surgeon |
| --- | --- | --- | --- | --- | --- | --- |
| All facilities | 3.8 | 1.8 | 12 | 563 | 62 | 302 |
| Min | 0.5 | - | 2 | 25 | 6 | 13 |
| Max | 9.0 | 4.0 | 29 | 2,924 | 353 | 2,924 |
| Private | 4.3 | 1.8 | 14 | 596 | 39 | 297 |
| Min | 0.5 | - | 2 | 25 | 6 | 13 |
| Max | 9.0 | 4.0 | 29 | 2,924 | 225 | 2,924 |
| Government | 2.2 | 2.2 | 6 | 473 | 128 | 314 |
| Min | 1.0 | - | 2 | 79 | 11 | 79 |
| Max | 6.0 | 3.0 | 9 | 809 | 353 | 705 |
| Rural | 3.0 | 1.8 | 7 | 343 | 79 | 207 |
| Min | 0.5 | - | 2 | 25 | 6 | 13 |
| Max | 9.0 | 3.0 | 20 | 809 | 353 | 705 |
| Urban | 4.6 | 1.8 | 16 | 762 | 47 | 387 |
| Min | 1.0 | - | 3 | 68 | 12 | 17 |
| Max | 8.0 | 4.0 | 29 | 2,924 | 225 | 2,924 |

A number of surgeons we interviewed reported feeling overworked, especially in having to manage outpatients during the day and accommodating surgical patients in the evenings.

> … eleven hours is the minimum hours that I spend here… Most of the surgeries I have done in the night…because I do not get time in the daytime. Whole day (nurses) will be walking around to see the patients…the same nurses have to go to surgery. To help me, so they get tired… also myself the same thing. (General surgeon, Assam, ACPT002)

The situation was exacerbated by general unavailability of skilled staff including assistant doctors and nurses. The lack of anaesthetists was a universal problem, prominently mentioned in the interviews, with a few also reflecting on the demoralising effect of shortages on their ability to do their work. Lacking dedicated anaesthetists, who would come on call, clearly added to available surgeons’ workloads. In some instances, surgeons mentioned multi-tasking, providing anaesthesia themselves or relying on nurse assistants.

> …sometimes we do not get anaesthetist in time so there we have to do by our self…sometimes nurses to give a spinal… (General surgeon, Arunachal Pradesh, APBPT002)

Notably, surgeons working on their own typically in smaller private facilities (<500 patients per annum, 10-20 bedded) lamented not having surgical colleagues in-house with whom they could discuss cases and procedures. This peer support was considered particularly critical for more complex cases involving excessive bleeding or suspected tumours. Clinical assistance was valued in such situations both for consultative purposes to guide medical decision making as well as for practical help performing more complex procedures.

> …anaesthesia is the biggest challenge here and another thing is the investigation facility. Sometimes… we can’t diagnose…because of lack of … CT scan, MRI, and endoscopy and sometimes we even have shortage of surgeons…(ideally) around 2-3 surgeons gather…discuss and…do the surgery properly…(But) we are left…alone to do one surgery which is very, very frustrating… (General surgeon, Arunachal Pradesh, APBPT002)

Some made do with existing nursing staff, often ones they had trained on the job. At other times, surgeons would consult more senior colleagues located in bigger cities.

> … I’m the only surgeon here and sometimes our medical superintendent, who is general practitioner…assist me in doing surgery and that becomes a little helpful for me. Otherwise I need to take the assistance of our sisters (nurses) who are also very well off now to assist during the surgeries. (General surgeon, Assam, ACPT002)

The above results provide an overall picture of the nature of workload and its effects on existing surgical provision, the implications of which are considered in the discussion.

### Referrals

Across surveyed facilities around 15% of patients requiring surgery are referred to other facilities. This level is slightly higher in rural compared to urban areas and from government compared to non-government facilities (Table 5).

**Table 5:** Surgical referrals

|  | Surgical referrals | Admissions | Surgeries | Referrals/admission | Referrals /patients needing surgery |
| --- | --- | --- | --- | --- | --- |
| Rural | 57 | 3,382 | 381 | 5.5% | 27% |
| Urban | 42 | 3,062 | 988 | 1.5% | 13% |
| Government | 94 | 3,807 | 387 | 7.1% | 25% |
| Private | 36 | 3,034 | 825 | 2.2% | 17% |
| Total | 48 | 3,197 | 733 | 3% | 19% |

Interviews revealed diverging demand and supply level motivators for referral to other facilities. Facility-level capacities were a principal factor prompting referrals by doctors, especially patients with co-morbidities or chronic disease. A few doctors, for instance, mentioned having to refer patients when anaesthetists were unavailable at short notice. At the facility level, a few doctors indicated diagnostic referrals to nearby private facilities for more advanced investigations like MRI or specialised procedures like endoscopy and laparoscopy. A doctor who held dual jobs at both a government and private facility, admitted having to routinely refer existing patients to the other private facility with functional infrastructure for laparoscopic procedures. Similarly, patients needing specialised surgery or complex cases that required more resources were referred to other facilities.

> …renal failure and … cardiac problem…so when they have complications, we need to refer and when their conditions is very poor… our hospital doesn’t have blood bank

> …abdominal traumas suppose someone is having liver injury, we need lots of blood so those kind of patients we cannot keep here…(Surgeon, Assam, ABPT002)

As also alluded to in the above quote, the survey revealed considerable variation in the adequacy of blood banking facilities. Except one respondent, a patient, who mentioned in-house blood banking facilities, most patients and surgical providers stated that patients needed to obtain blood from external sources. Family donations or blood banks in other locations were often used. A doctor in a private mission hospital (19 beds, 1 functioning Operation Theatre, approx. 1032 admissions per annum, 2 doctors) mentioned that they would either stabilise those likely to need blood transfusions or rely on their experience before proceeding with a surgery, using patient haemoglobin levels as a yardstick. For such facilities in small towns, other infrastructure priorities and regulatory factors disincentivised investment in blood banking.

> The licences [for a] blood bank are very difficult to get. We need pathologists…dedicated for the blood bank…there is lot of norms …staffing patterns, the room, the chairs… A lot of things are required…These will cost lots of money–we cannot keep as a priority a blood bank …We ask for the blood but if the haemoglobin is around nine or ten I’ll definitely go ahead because getting blood is a [big problem]. (General surgeon, Assam, ACPT002)

Along with limited equipment, this lack of surgically trained staff was a contributing factor for less routine use of minimally invasive laparoscopic techniques. Despite interviewed surgeons’ amenability to laparoscopy, some pointed out that they were using it mainly for elective rather than acute cases requiring advanced laparoscopic capability, lacking in their facilities. This was because of both affordability for patients as well as surgeons’ avoiding potentially more complex cases via laparoscopy, which they had limited training and experience of. Specifically, some surgeons acknowledged clinical and economic benefits of laparoscopy including quicker recovery time and less pain, which given their patient profile of less well-off populations was likely to be a preference for patients as well.

Interviews with medical staff indicated that record-keeping varied greatly between facilities. Some facilities digitised records whereas others kept written registers. In general, public hospital data was regularly sent to government health information management systems.

### Treatment seeking, cost, and pathways of care

Patients in our sample were generally satisfied with the care received, mainly because they had their ailments addressed. A range of considerations were noted in patients’ descriptions of care experiences—doctor and medical staff manner, patient’s prior experience and hence faith in using a certain facility, and for one user in Manipur, a shared dialect with medical staff which improved communication and comfort level. Good interactions with staff as well as clear communication from medical staff reassured patients were important for patient satisfaction. The following respondent who had their pain symptoms persist and then sought treatment at a private facility reflected on their earlier experience at a government hospital.

> I was abruptly discharged… what I mean the doctor will come for the round-’she is fine-discharged’… so I was a bit disappointed… that they discharged me because I do not know what my condition is … because the pain is still there… I wish the doctors can be more reassuring then they can give us more advice. (Female patient, 40-45 years, private facility, Manipur MCPT001PTE)

Interviewed patients generally reported self-treating when symptoms, typically abdominal pain, first emerged. Getting medicines from local pharmacies was the preferred option for most patients. Patients in our sample generally preferred consulting medical doctors over traditional practitioners, often because of faith in formal care as well as symptoms like acute abdominal pain needing emergency or obstetric care. Shorter distances to facilities, reputation of doctor/facility, and for some, lower costs of services were prominent factors affecting patients’ initial choice of facility. A few responses also indicated an acceptance of poor roads and traffic conditions, without noting these as factors affecting access to facilities. Convenience and cost emerged as prominent drivers for seeking services from multiple providers resulting in slighter longer pathways to getting treated.

> I went to the other hospital because it’s nearby and my case was an emergency. But I got shifted to this hospital because it is cheaper here. I took doctor’s reference to come here. (Female patient, 30-35 years, private facility, ABPT002)

The facility survey revealed that the cost of essential surgery varies across health facilities but is on average between 5 and 8 times higher in non-governments compared to government hospitals. Average charges in non-government hospitals constitute between 37% (C-section) and 47% (fracture repair) of annual state GDP per capita (Table 6).

**Table 6:**
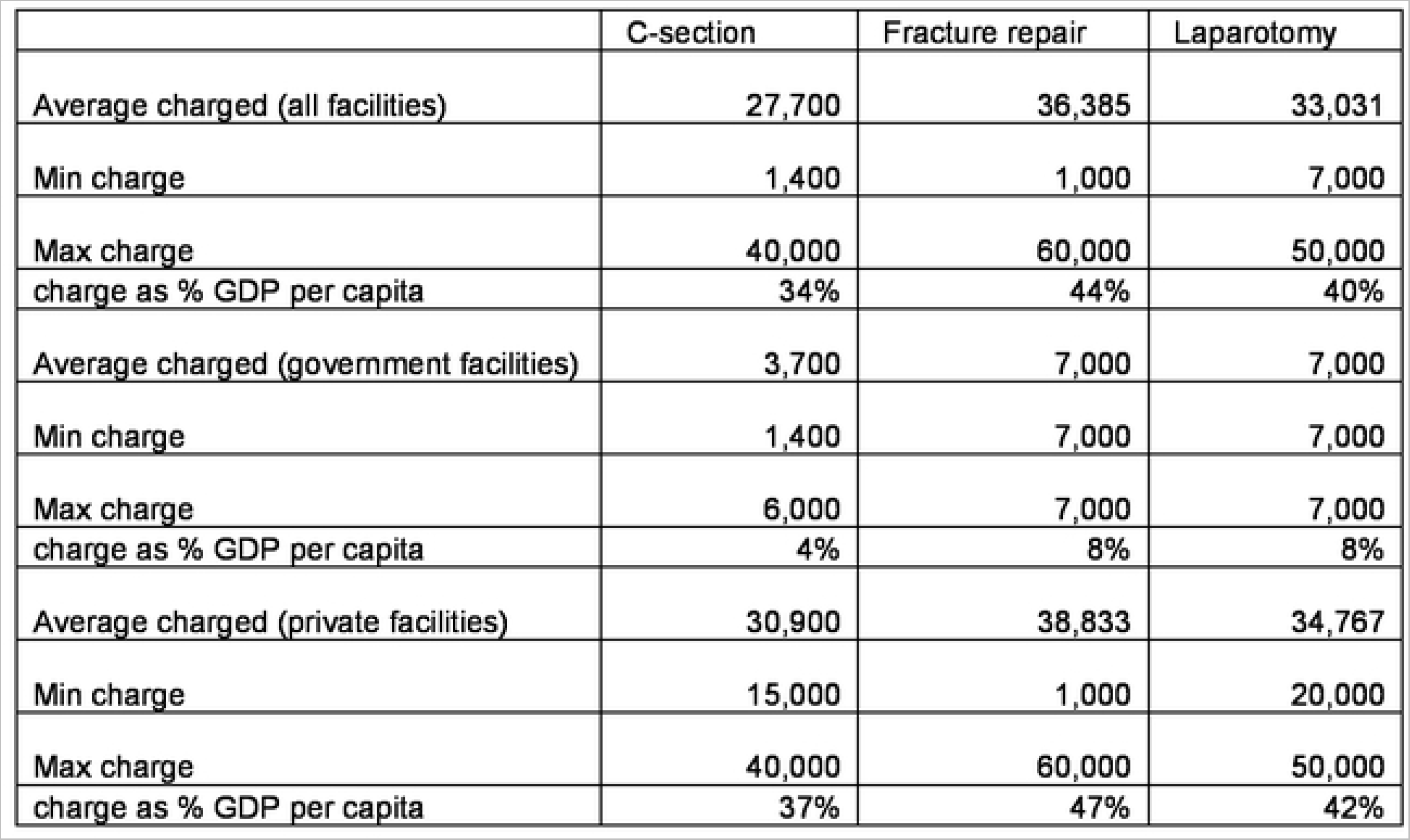
Facility charges of essential surgery (Rupees)

## Discussion

The quantitative and qualitative results collectively provided critical insights into surgical care capacities in NE India, current practices, and the readiness for adoption of new surgical innovation.

Abdominal conditions constituted a large portion of the overall surgeries across public and private facilities in the region. The prominence of caesarean section was evident, on the positive side signalling the availability of emergency obstetric care, but equally a more portentous trend towards avoidable procedures (WHO 2021). This was partly reflected in the higher uptake of surgery among women, which could indicate the magnitude of obstetric health needs in the region while also raising questions about men’s health seeking behaviours.

Facility functionality and being better equipped, was an enabler for surgical delivery, although other considerations may also explain lower caseloads in some facilities. An accompanying paper, for example, suggests that location of facility and distance are critical factors in determining use of facilities, suggesting lower take-up by users even in cases where surgical resources may be available (Ensor et al, 2021). Results demonstrated variable workloads for surgical providers—interviewees generally reporting excessive workloads although the facility survey also revealed smaller surgical volumes at certain facilities. Interviews more clearly revealed the pressures that excess workloads placed on existing personnel both surgeons and nurses. Scarce peer support for complex cases and a reliance on nursing staff created physical and cognitive demands on surgeons. More broadly speaking, skewed workloads whether high or low, have critical implications for provider efficiency, patient safety, and care quality. High workloads, for instance, can overburden staff, affecting their motivations and performance, and in some instances leading to preventable clinical errors and sub-optimal treatment (Michtalik et al 2013, Yu et al 2016). Conversely, low surgical volumes at some facilities may limit opportunities for staff to get sufficient practice, leading to adverse patient outcomes. For instance, Mikeljevic and colleagues (2003) reported lower survival rates among breast cancer patients treated by surgeons with lower workloads. More broadly, skewed workloads across facilities and regions are indicators of uneven surgical delivery, with lack of uniformity in care quality and facility performance.

There was some evidence of task-shifting, in non-specialist nursing staff assisting doctors on surgical procedures or surgeons’ taking on anaesthetic tasks or supervising, as has occurred elsewhere in the US, Western Europe, Sub-Saharan Africa (Federspiel et al 2015). In Sierra Leone, task shifting involves training non-surgeons to perform certain operations rather than nursing staff undertaking dual roles. Mavalankar and Sriram (2009) view task-shifting to mid-level providers as a promising strategy given personnel and training shortages for specialised anaesthetic care in South Asia. According to the authors, in addition to better staff retention amongst mid-level providers, effective training and quality assurance measures can ensure the cost-effectiveness of safety of task shifting approaches in South Asia. Anaesthesia is a vital element of emergency obstetric care, needed to tackle pregnancy related complications, especially prominent in South Asia and accounting for high rates of maternal mortality.

Additional to personnel and infrastructure shortages, shortfalls in-house blood banking facilities were evident, adding time and costs to patients’ treatment pathways, as blood supplies had to often be externally sourced or replaced through family donation. Whilst the shortages in blood supplies globally as well as in India are well known due to a combination of demand and supply level factors, our results also revealed structural disincentives including tedious licensing norms for in house storage discouraging active investment in blood banking facilities (Barnes et al 2022, Mammen 2022). Prominently, there was amenability to greater use of minimally invasive laparoscopic techniques, which are routinely used in more developed health systems given relative cost, time, and efficiency advantages over open surgery (Chao et al 2016). Clearly, however, equipment and training incapacities were acknowledged signalling the direction of possible policy action.

Survey results revealed high rates of referral for some (particularly government) hospitals to other facilities – often because surgeons or equipment were not available. Interviews with surgeons at private clinics confirmed facility-level infrastructural shortcomings or personal incapacity as factors prompting referrals to external facilities. Whilst case complexity is often legitimate grounds for referral to higher levels of care, our results suggest that avoidable treatment delays were occurring due to a lack of basic diagnostic infrastructure in several mid-range facilities. In our results, the one striking instance of a surgeon transferring public patients to private facilities raises concerns about affordability for patients and equity within the system, especially as we found that for users, cost of care often functioned as a driver for self-referral to cheaper often public facilities for procedures. Essential procedures were expensive at non-government facilities which has implications for affordability and equity of access within the system. More broadly, this example provides further evidence on the factors motivating referral to private facilities (Michael et al 2022). In addition to doctors’ personal interest and circumstance as motivators in some contexts, our work shows that doctors’ assessment of patient’s financial ability and preferences also factoring into referral to better equipped (with investigation technology) or cheaper (public facilities, or electing for more technologically advanced procedures (eg. laparoscopy).

## Study Limitations

Admittedly, the providers and patients in the sample of interviewees captures a limited set of views from certain districts. Yet, these accounts provide rich detail on evidence uncovered through the facility survey, providing a comprehensive overview of surgical delivery in the region. Moreover, while included patients were generally female and often being treated for abdominal conditions, the interviews nevertheless represent a prominent population and category of surgical need in the country. Difficulties accessing information from the entire universe of surgical facilities meant that certain categories namely military and certain missionary facilities, important providers of care in the region, were excluded from the results. However, the sites are broadly representative of the general capacity for surgical care and experiences of those privileged enough to be able to access surgical services. In tandem, the quantitative and qualitative assessments provide a picture of surgical care in Northeast India.

## Conclusion and recommendations

Service availability, represented in functioning facilities and adequately trained staff, is vital for surgical access. In low resource settings as in Northeast India, informal arrangements for task-shifting and referral function as common strategies for surgical providers to cope with surgical demand. The motivations for referral often stem from facility incapacity in certain specialised areas of care, such as anaesthesia and advanced diagnostics. Such shortages call for structural changes to facilitate procurement and training in priority specialties. Innovations in technology, and management practices are needed. Digital technologies are becoming available that offer affordable and efficient ways to enhance surgical training through remote proctoring, and frugal innovations, such as gasless laparoscopy, which may allow patients in rural settings to benefit from minimally invasive procedures. On the organisational side, better coordination of referral among existing facilities could go some way to ensure more integrated and efficient services. On staffing, a system wide approach is needed to ensure a coordinated response to addressing scarcities in human resources and skill-mix. Finally, while health system level levers are vital for enhancing surgical delivery, wider political, economic, and structural change to ensure adequate prioritisation and financial investment will be crucial to address other components of access to care including economic and geographic aspects.

## Ethics

Ethical approval was obtained from Sigma Research and Consulting (India) and the University of Leeds School of Medicine Research Ethics Committee (U.K.).

## Data Availability

All relevant data are within the manuscript and its Supporting Information files.

## Acknowledgements

Lanuakum Aier, Catriona Child, ThepfuchanuoKire, Atshele Venuh, Benrilo Shitiri, Keduokuolie Pienyu, Lungreihungbe B. Riame, Seyievor Yhome.

